# A Geospatial Analysis of salmonellosis and its association with socioeconomic status in Texas

**DOI:** 10.1101/2021.05.21.21257607

**Authors:** Anand Gourishankar

## Abstract

**Objective:** The study’s objective was to find the association between salmonellosis and socioeconomic status (SES) in hot spot areas and statewide counties.

**Design:** A retrospective cohort study.

**Setting:** The data was recorded regarding salmonellosis in 2017 from the Texas surveillance database. It included assessment of hot spot analysis and SES association with salmonellosis at the county level.

**Participants:** Patients with salmonellosis of all age groups in Texas.

**Results:** There were a total of 5113 salmonelloses from 254 counties with an unadjusted crude rate of 18 per 100,000 Person-year. Nine SES risk factors in the hot spot counties were as follows: low values of the severe housing problem, unemployment, African American, and high values of college education, social association rate, fast food/full-service restaurant use, Hispanic, and senior low access-to-store (*P* < 0.05). A 12% difference existed between local health departments in hot (25%) and cold spot (37%) counties (*χ*^*2*^ [1, *n* = 108] = 0.5, *P* = 0.81). Statewide independent risk factors were severe housing problem (IRR = 1.1; CI:1.05-1.14), social association rate (IRR = 0.89; CI:0.87-0.92), college education (IRR = 1.05; CI: 1.04-1.07), and non-Hispanic senior local access-to-store (IRR = 1.98; CI: 1.26-3.11). The severe housing problem predicted zero occurrences of infection in a county (OR = 0.51; CI: 0.28-0.95).

**Conclusions:** Disparity exists in salmonellosis and socioeconomic status. Attention to unmet needs will decrease salmonellosis. Severe housing problem is a notable risk.

## BACKGROUND

Salmonellosis is one of the leading foodborne illnesses in the United States. Salmonellosis (NTS; nontyphoidal salmonellosis) is defined as a clinical illness that results from infection by serotypes of Salmonella other than Typhi or Paratyphi (A/B/C) that causes Typhoid fever. Ranked from the fewest to the greatest number of Salmonella all-type outbreaks, Texas ranked 15^th^ among 50 US states (1999-2017) before dropping to 36^th^ in 2019. In the five years from 2011-2015, the average number of salmonellosis cases reported in Texas was 5,205 cases per year (range: 4,946 to 5,727). Salmonellosis accounted for 31% of all foodborne illnesses.(1) Salmonellosis risk varies by serotypes. Also, socioeconomic status (SES) influenced the risk of exposure to a specific pathogen (1, 2). A systematic review of four foodborne-illness pathogens, including Salmonella, showed that higher SES, higher education, unemployment, poverty, urban area, and lower deprivation were associated with higher incidence.(1) Geographic Information System (GIS) and Spatial Analysis have expanded into the science of spatial epidemiology (3-5). Area-based socioeconomic status (ABSES) such as levels at the county, census tract, or block group analysis may provide information about transmission and the opportunity to prevent infections (6). The population-based ecological studies are limited in number and not generalizable, as described further here. A study in Canada, using county-specific SES and disease rate, found that salmonellosis was associated with age < 5 years, non-white race, and poverty (7). A study that used block group-level found that decreasing years of education was associated with reduced salmonella infection (8). A previous census tract-level study in Connecticut found an association between high SES and NTS in persons ≥ 5 years of age and some serotypes (9). A county-area level nonspatial analysis for SES found unemployment, Black, Hispanic, and Latino ethnicities were associated with NTS (10). Currently, there is no spatial analysis of area-based (county-level) SES and NTS in Texas.

A county-level assessment will provide new insights for community interventions to reduce the infection rate and community health service disparities. The objective of the current study was to analyze the relationship between nontyphoidal salmonella infections and county-level SES in the hot spot areas and across all counties within the state of Texas.

## METHODS

The estimated population for each Texas county was derived from the US Bureau of Census. County-level SES was derived from The County Health Rankings & Roadmaps program and Food Environment Atlas data (Table 1). For this study, the county-level NTS (2017) across Texas were retrieved from the Texas department of health (www.dshs.texas.gov). All the data were joined to administrative boundary shapefile of census tracts obtained from the TIGER/Line database (www.census.gov) using ArcGIS Pro 2.5 (ESRI Inc., Redlands, CA, US).

**Table 1.**
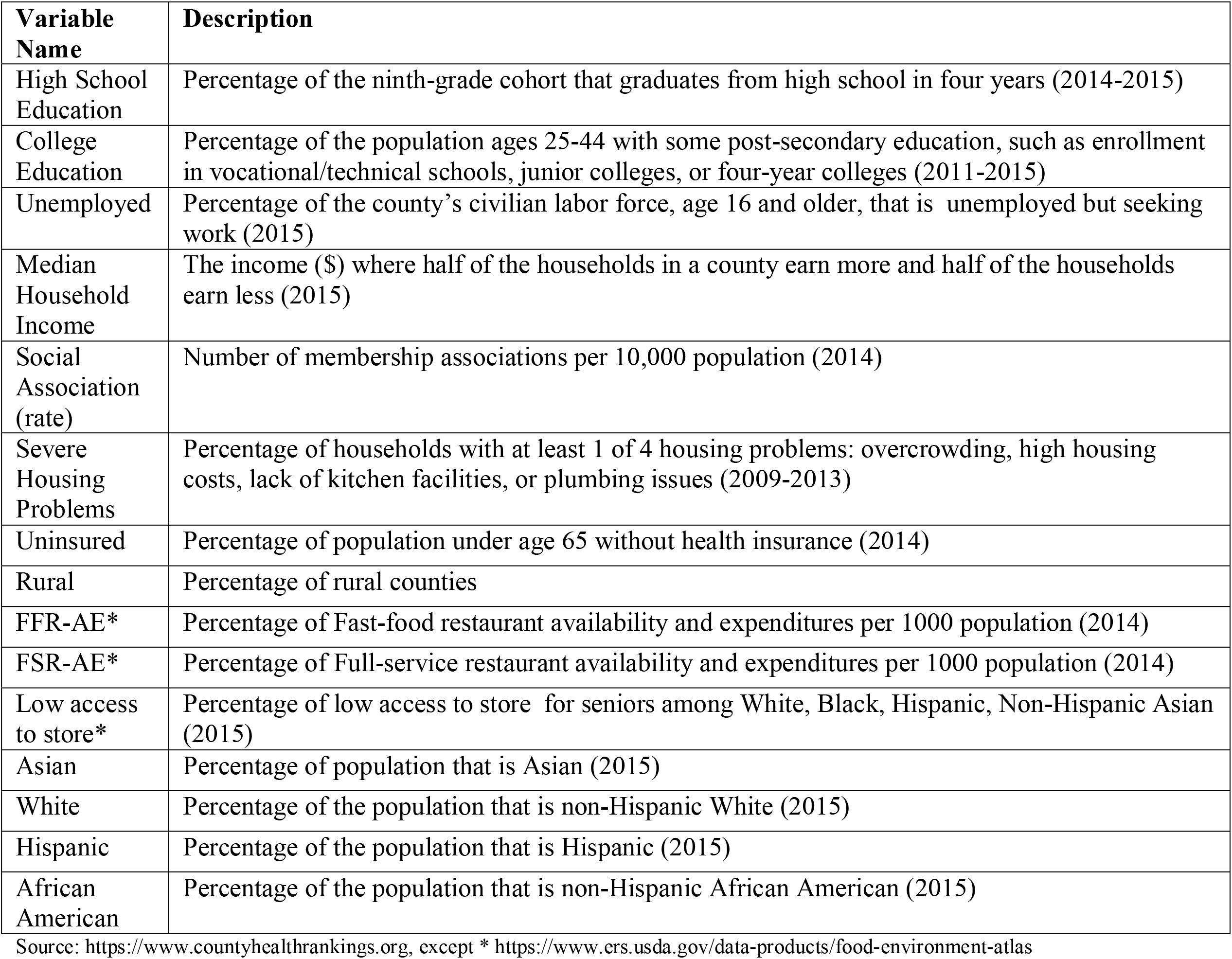
Explanatory variables: definitions and sources.

### Statistical analysis

Analysis of the total population and NTS counts included descriptive statistics. The mean (Standard deviation) and Median (Interquartile range) were calculated for continuous SES variables. Data distributions of SES variables determined the inferential statistics (Student t-test or Wilcox test). The chi-squared statistics compared the proportions of local health departments of Texas. The spatial Empirical Bayes (EB) smoothing method reduced the random variation of infection rates. It accounted for unstable incident rates in areas with small populations (second-order queen contiguity weights using R software 3.6, Vienna, Austria) (11, 12).

### Spatial analysis

A choropleth map was created with the Texas county base map and Jenks optimization classification method in ArcGIS Pro. Choropleth will show a large-scale variation of salmonellosis incidence rate per 100,000 population (normalized) in each county. Moran’s Index represented global clustering (spatial autocorrelation) of counties with high or low salmonellosis across Texas. Moran’s Index measures proximity by inverse distance weights - an Index that combines two measures of attribute similarity and location proximity into a single index.(7) Moran’s Index is considered statistically significant with a high z-score and *P* ≤ 0.05. Next, local clustering of the disease investigates the spatial variations and spatial associations. Therefore, optimized hot spot analysis (extension of Getis-Ord Gi* statistics) identifies hot and cold spots by comparing individual high or low values (NTS counts) with surrounding other high or low values, respectively. The optimized hot spot analysis automatically selects an appropriate analysis scale and adjusts results for multiple testing and spatial dependence. A large positive z-score that is statistically significant (*P* ≤ 0.05) detects hot spots. Similarly, cold spots have a largely negative and statistically significant z-score. Choropleth map can display the hot/cold spot z-scores with confidence intervals in red and blue colors.

### Modeling approach

Multiple linear regression was associated with multicollinearity and residual autocorrelation. Also, the NTS counts were skewed. The overdispersion and 32 counties with excessive zero-infections had limited Poisson regression model fit. Therefore, we applied the zero-inflation negative binomial regression model (Zinb). The univariable screening included the Zinb regression of the sixteen variables (Table 1).

Spearman’s correlation (Rho > 70) test applied to 16 variables. Variance inflation factor detected multicollinearity. Forward regression was carried out using *P* < 0.05 to include variables until the final model. The Chi-squared test on the difference of log-likelihoods and the Vuong test were used for the final model’s diagnostics. The demonstration of absent residual clustering (autocorrelation-hotspot) using ArcGIS Pro will also confirm the model fit.

## RESULTS

In 2017, there were 5,113 cases in 254 Texas counties. The average unadjusted county’s crude incidence rate (IR) of salmonellosis in Texas was 18 per 100,000 Person-year. The median IR (per 100,000) was 20 (IQR: 11 & 36). Fourteen percent (35/254) of counties had zero salmonelloses. The annual county population for the year 2017 ranged from 80 to 4,629,700 (median = 18934; IQR: 6,975 & 52,020). Choropleth map illustrated the county-level comparison of crude IR and Empiric Bayesian smoothed rate. Despite EB smoothing of unstable rates, visual inspection of both maps showed (large scale) almost similar variations in Texas (Figure1).

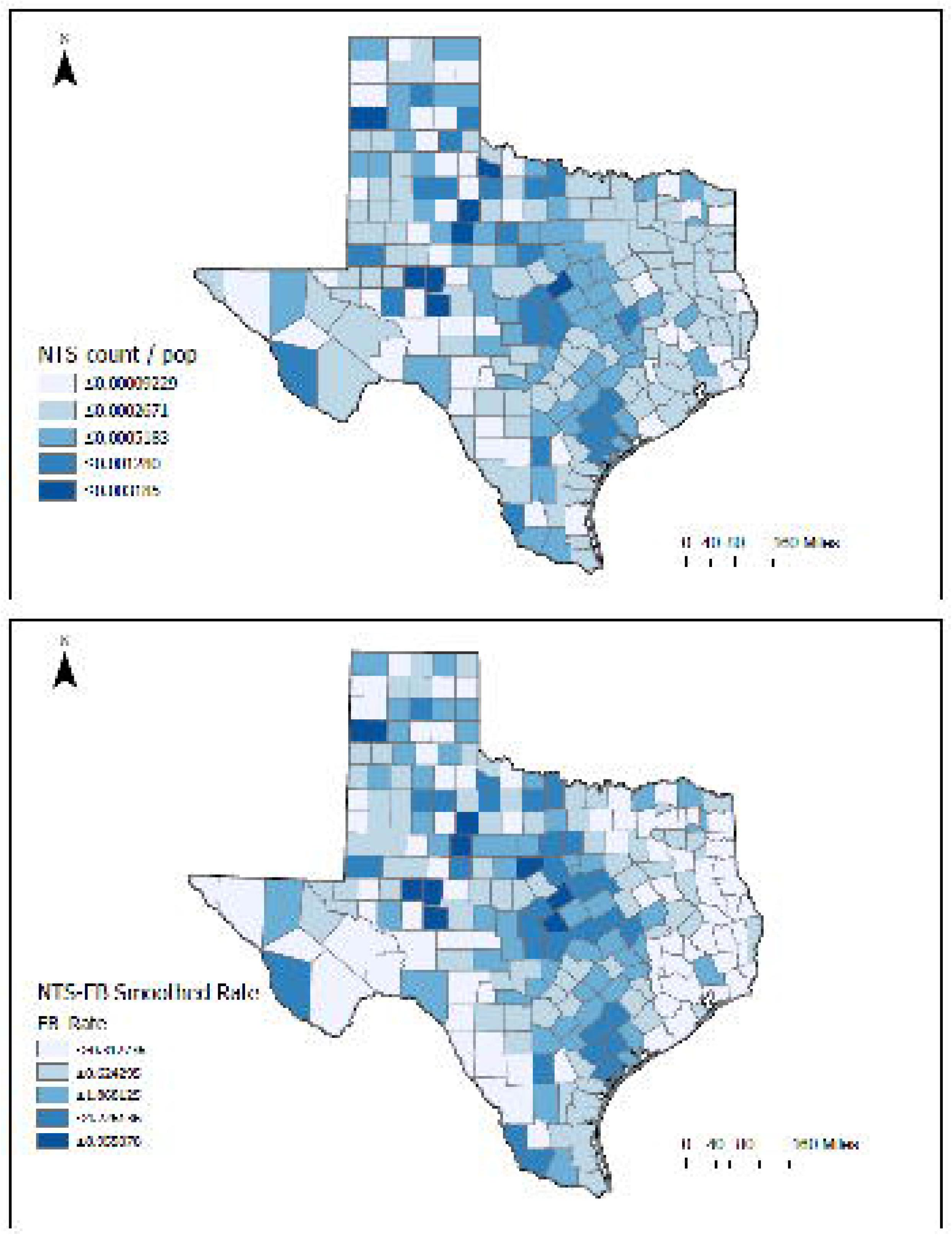

Geographic clustering of the EB smoothed rate was high in central Texas (red) compared to low rates (blue) in east Texas (Figure 2A). Hot spot corresponds to public health regions 2|3 and 7, and cold spot corresponds to public health regions 4|5N and 6|5S (Figure 2B). The global spatial clustering measure, Moran’s Index was 0.1 (*P* < 0.001), which explains that the salmonellosis cluster pattern (hotspot) was unlikely due to random chance. In general, this study found an association between salmonellosis and nine SES indicators (Table 2).

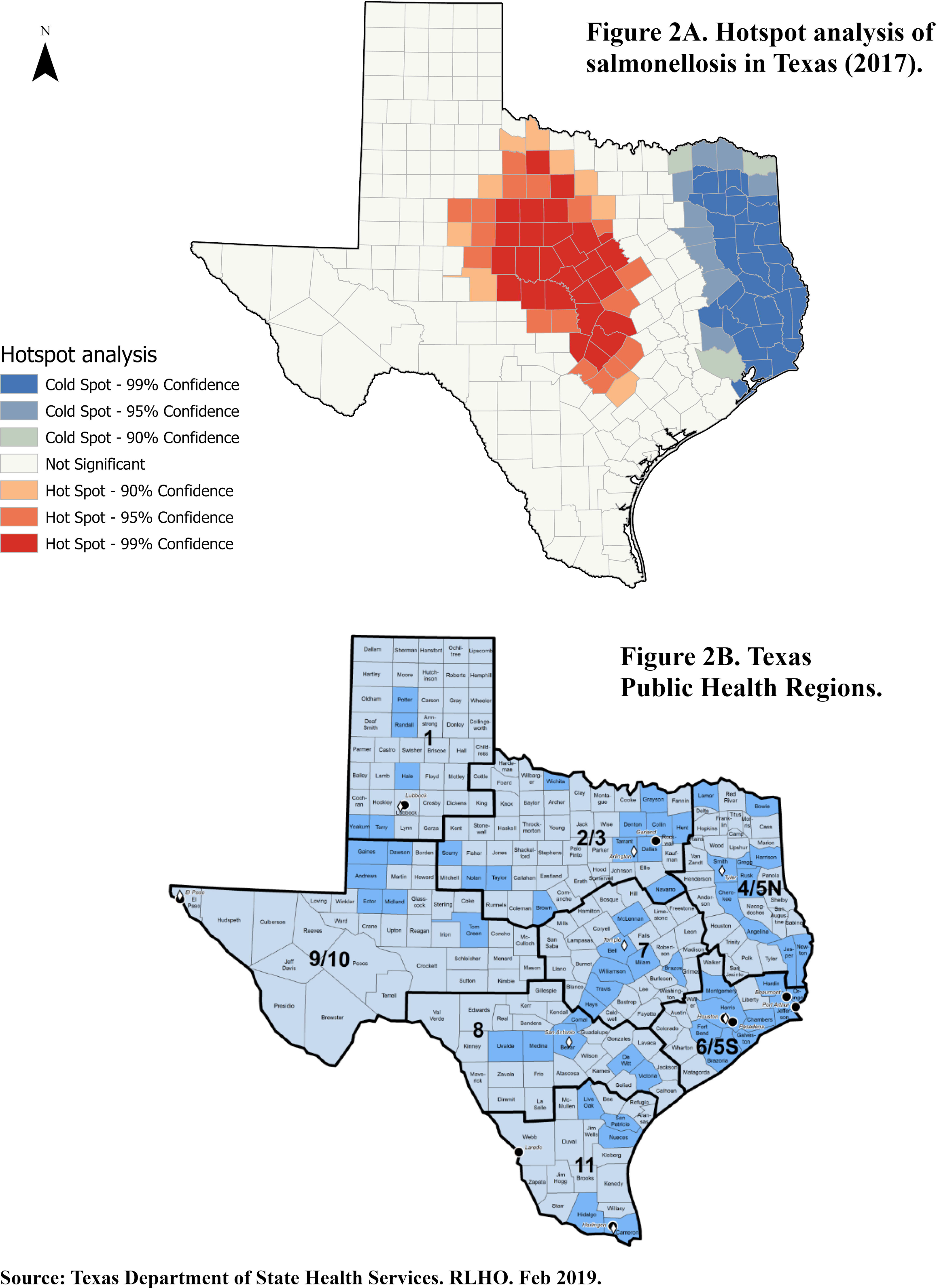

**Table 2:**
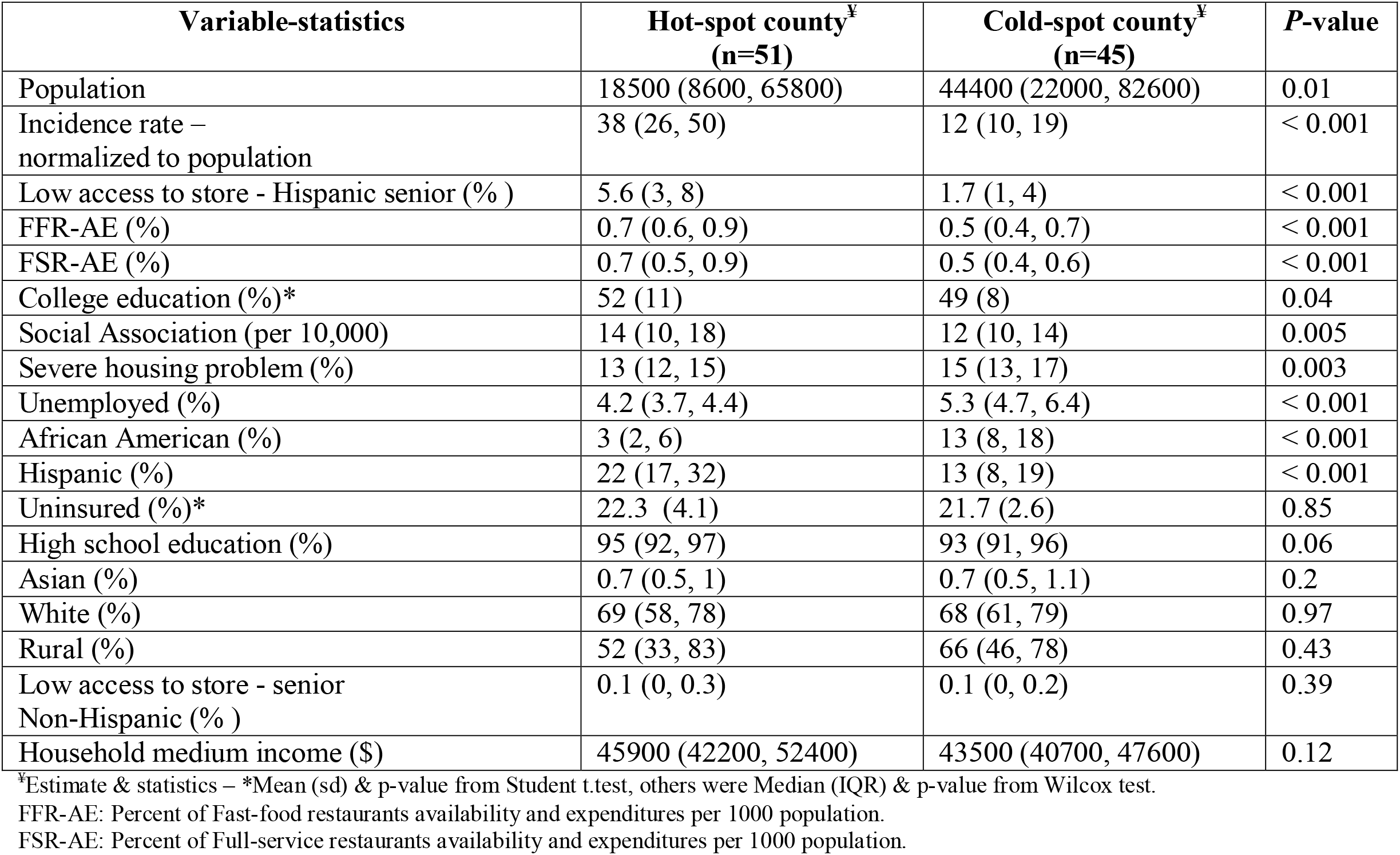
Comparison of variables between counties identified by hotspot analysis.

When comparing hot and cold spot counties, the following SES indicators were significant (*p* < 0.05): ‘low access-to-store in senior Hispanic’, Hispanic, and African American ethnicities. Hispanics were 22% compared to 3% of African Americans in hot spot counties. On the contrary, the proportions of White were higher than other races in both hot and cold spot counties without statistical significance. The ‘social association rate’ (high means healthy) was high in the hot spot, and ‘severe housing problem’ (low means healthy) was high in the cold spot. The study found high proportions of fast-food and full-service restaurant availability and expenditures in hot spot counties. Median income was not different between the groups, but high unemployment was in cold spot counties. There was a higher percentage of college education in the hot spot than the cold spot (52% vs. 49%). There were no differences in those with high school education. Furthermore, there was a disproportionate distribution of local and city health departments among the 11 public health regions in Texas. Higher values of salmonellosis (hot spot) was noted in the regions with lower number of health departments (Hot spot = 25%, Cold spot = 37%, *χ*^*2*^ [1, *n* = 108] = 0.5, *P* = 0.81, Figure 2A & B).

In the global multivariable model, among the 16 independent variables, the final model included only four variables. These variables were ‘severe housing problem’ (%), ‘social association rate,’ college education (%), and percentage of ‘low access-to-stores in non-Hispanic Asian seniors’ (Table 3). Estimates (IRR or OR) resulted from one unit increase in the final variable, provided the other variables were held constant in the model. As such, for ‘low-access-to-store of non-Hispanic Asian seniors,’ ‘severe housing problem,’ and college education, the rate of salmonellosis would be expected to increase by a factor of 1.98, 1.1, and 1.05, respectively. By contrast, if a county had an increase in ‘social association rate’ by one unit, the salmonellosis rate would be expected to decrease by a factor of 0.89. The ‘severe housing problem’ predicted zero occurrences of salmonellosis. If the percentage of ‘severe housing problem’ increased by one unit, there were 51% odds that a county would have “zero” salmonellosis. Thus, the higher the housing problem, the more likely the county had zero infection rate (Zero-inflation model, Table 3).

**Table 3.**
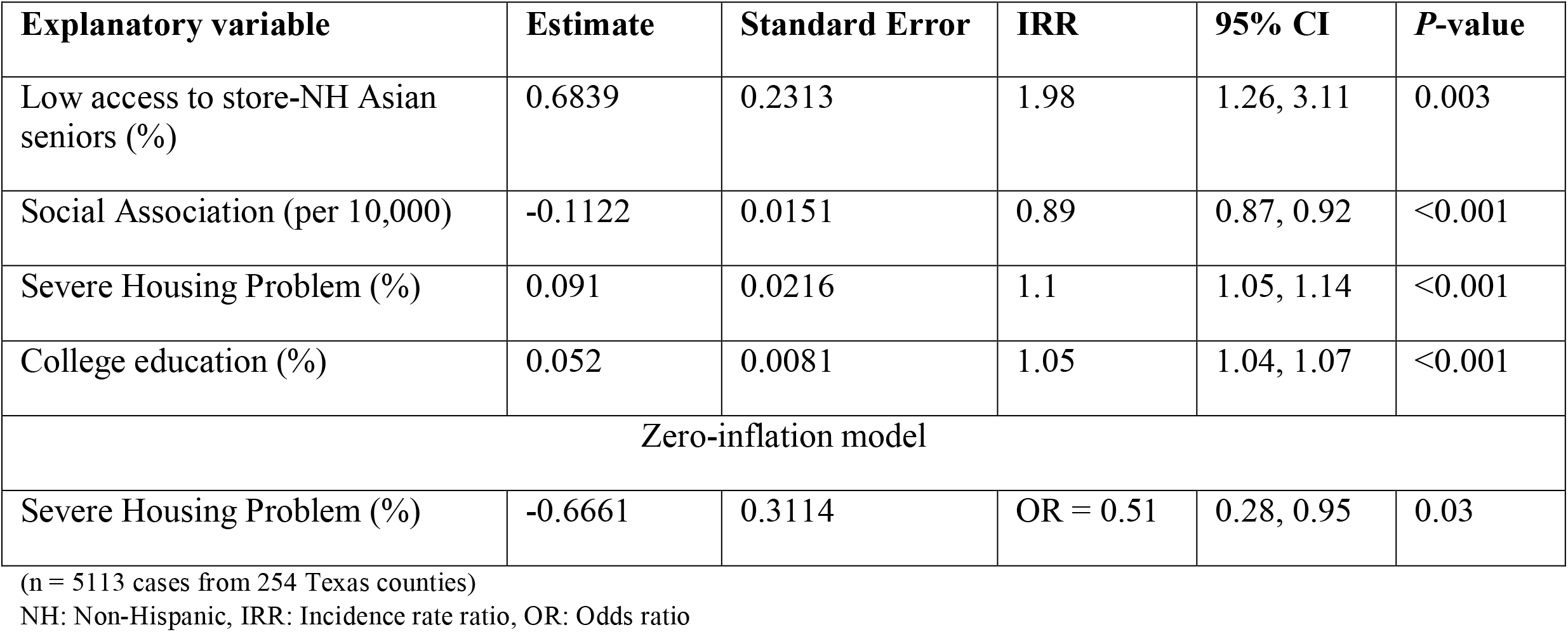
Results of the final multiple variable zero-inflation negative binomial regression model.

## DISCUSSION

Salmonellosis of Texas in 2017 showed disparities in regional clustering of cases, public health regions, and socioeconomic status indicators at the county area level. Optimized hot spot and global Moran’s Index method identified 51 counties in the central part of Texas with a high incidence rate. The higher incidence of cases in the central part of Texas can be attributed to differences in the availability of public health regions and SES indicators. The central Texas (Public Health Regions 2/3, and 7) presented with high disease clusters (statistically significant hot spots, Figure 2 A & B). Although no statistical significance, a gap of 12% in the presence of local health department between cold and hot spot regions is meaningful. Expansion of health services and expenditure may reduce salmonella incidence (13, 14). There was a statistically significant association between nine SES indicators and salmonellosis in the hot and cold spot counties in bivariate analyses. Based on this study, a combination of four SES indicators of ‘severe housing problem’ (%), social association rate, college education (%), and ‘low access to store’ in non-Hispanic Asian seniors (%) could explain the variability in the occurrence of salmonellosis in Texas.

At county-level GIS analysis, this study is the first one to demonstrate hotspot and SES association regarding salmonellosis (broad, not species-specific). Previous studies had attempted a similar effort to analyze SES at different area-based levels; GIS analysis at the census tract-level showed the association of age and high SES in certain Salmonella species (9). The block-level analysis had demonstrated the association of decreasing years of education with a decrease in salmonella infection (8). Both these two studies showed a general trend without hot spot analyses. Although the current study did not analyze various serotypes, it is unclear why specific serotypes affect high SES. Varga et. al found *Salmonella enteritidis* area-level hot spot clustering with SES indicators (income, visible minority, number of children/family) (7). The hot spot analysis method was different between this study and Vargas et al. Understanding these differences includes the interplay of various factors; physical, biological, behavioral, cultural, health services utilization, SES, and environmental factors. The current study results were consistent with some of the results from these studies. However, they could expand on analyzing various county-level serotypes in the future.

A higher percentage of a college education was associated with high clusters of salmonellosis in this study. Most, but not all, studies have shown associations between high educational attainment and infection (8, 10, 15). It is postulated that higher education increases awareness of food safety labels (16). By contrast, the decreased incidence was reported in low SES (low education and income) due to the better practice of hand hygiene, less eating of risky food, and better food storage (17-19). Other explanations for higher education and infection associations are greater access to healthcare, health-seeking behavior, pet ownership, and eating fresh produce, raw or uncooked food (20, 21). Unemployment has a protective effect on salmonellosis. This factor by itself or concurrent with a lower education level can explain this observation. An Italian study by Borgnolo et. al found higher non-Typhi salmonella infection rates in children whose fathers were either unemployed or working in non-blue-collar jobs (22). SES indicators, economic status (income), and higher educational attainment are intertwined, manifest as a differential effect of SES in salmonellosis. By contrast, this study did not show a salmonellosis association with median household income consistent with previous research (15). Hence, the current study underscores the several explanations that interplay between education and economic status.

In a study by Lay et. al (23) African Americans had a higher incidence of salmonellosis, whereas this study found a higher percentage of African Americans in cold-spot counties. In Younus et al. (8), they found no association with ethnicity and salmonellosis, but our study found that the Hispanics were higher in hot spot counties, consistent with Arshad et al. (24). Ethnicity may be a function of individual risk factors and pathogen-specific (ecological effect of serotypes and SES) (9). Although disparities exist, it is an unclear association between foodborne pathogens and ethnicities (25). The disparity in salmonellosis among ethnicities can arise from gentrification and housing segregation. Thus, exposing the population segment to any number of high-risk SES indicators noted in this study. Although the Hispanic ethnicity emerged significant, demographic factors, behavioral or cultural, and other individual risk factors may affect these associations.

The other highlight of this study was that the fast-food and full-service restaurant utilization among seniors in the county was associated with salmonellosis (seniors are a known, high-risk age group). Darcey et al. and Signs et al. have found differences in SES with health code violations and food safety in retails, respectively (26, 27). Appling et al. reported the risk of salmonella infection and violations in the restaurant (28). Although the differences between hot-cold counties on restaurant use in this study were small but statistically significant, further studies can strengthen this study’s findings. Furthermore, low SES communities are more likely to visit fast-food restaurants (29, 30). Fast-food and full-service restaurant availability and expenditures can be associated with economic disadvantages such as poverty, unemployment, or low educational attainment. Despite food safety measures by agencies and food education, SES indicators are the significant determinants for salmonellosis. The ‘low access to stores for seniors’ had divergent results for Hispanic compared to non-Hispanic. There may be a bias due to a higher percentage of Hispanics in the hotspot counties.

The ‘social association rate’ is a powerful predictor of health status (positive perception and behavior of health)(31). Although the ‘social association’ is a ‘rate,’ limited by self-reporting of local entities, it measures vital health-related memberships, such as fitness centers, sports organizations, religious organizations, civic and business organizations (32). Social networking and community improvement (social capital) support the belief that if individuals are not isolated and have strong social networks, they make healthy choices.

Based on this study, ‘severe housing problem’ (a measurement of the percentage of lack of kitchen or plumbing facilities, overcrowding, and severely cost-burdened) demonstrated substantial influence in salmonellosis. It is the only indicator that predicted the “zero” occurrence of salmonellosis infection. Although it appears protective, it underscores the magnitude and strength of adverse societal problems (33). ‘Severe housing problem’ also reflects reduced food access, poverty, positive food storage, and likely underreporting and less access to health care (6, 13, 34). Adequate housing (a proxy of high SES) prevents harmful exposures and provides a sense of safety, contributing to health. The current study supports allocating resources and services to home environment assessments, indoor pest management, grants for community development, housing, and inclusionary zoning and housing policies (35).

### Limitations

SES is challenging as it lacks a single metric and SES index. Also, measurements vary among studies for a single variable. However, Jouve et. al had described the inherent issue of a complex interaction between SES and the outcome of interest, a function of differential exposure and differential vulnerability (36). Under-reporting (decreased case ascertainment) due to passive surveillance of salmonellosis reduces the true incidence. Finally, ecological analyses do not assess confounding, and “ecological fallacy” is inevitable. Although multiple regression addresses confounders, the final model is susceptible to misspecification. The study’s strength includes group-level analysis accounting for both individual and community level SES, rigorous hotspot analysis, no missing data, and modeling at the local and global levels of SES. County-level analysis can miss individual-level variation, whereas it provides information for directing policy and resources to the community.

## Conclusions

The regional disparity in salmonellosis demands better research, improved capacity, and an effort for surveillance to identify actual infection rates and the allocation of resources. The weight of the evidence suggests improving socioeconomic status indicators, and access to health services can reduce salmonellosis in Texas and across the United States.

## Data Availability

No data available

## Data availability

The data were derived from the following resources available in the public domain:

1. Texas department of health; www.dshs.texas.gov
2. RWJ Foundation Program. County Health Rankings and Roadmaps 2020; https://www.countyhealthrankings.org/.
3. Food environment atlas data; https://www.ers.usda.gov/data-products/food-environment-atlas.

## Notes

### Competing Interest Statement

The authors have declared no competing interest.

### Funding Statement

No Funding

### Author Declarations

No IRB is required for this study.

